# Automated pupillometry to detect residual consciousness in acute brain injury

**DOI:** 10.1101/2024.02.03.24302210

**Authors:** Marwan H. Othman, Markus Harboe Olsen, Karen Irgens Tanderup Hansen, Moshgan Amir, Helene Ravnholt Jensen, Benjamin Nyholm, Kirsten Møller, Jesper Kjærgaard, Daniel Kondziella

**Author notes:** Corresponding author:* Daniel Kondziella, MD, Dr Philos., MSc, FEBN, Department of Neurology, Copenhagen University Hospital - Rigshospitalet, Inge Lehmanns Vej 8, DK-2100 Copenhagen, &. **Details page** The authors confirm the following: The manuscript complies with all instructions to authors; authorship requirements have been met; the final manuscript was approved by all authors; this manuscript has not been published elsewhere (but it is submitted to a preprint server, i.e. MedRxiv) and is not under consideration by another journal; ethical guidelines were adhered to and we indicate ethical approvals (IRB) and use of informed consent, and we confirm the use of an appropriate reporting checklist. **Author Contributions:** Drs Othman and Kondziella had full access to all the data in the study and take responsibility for the integrity of the data and the accuracy of the data analysis. **Concept and design:** Daniel Kondziella, Marwan H. Othman. **Drafting of the manuscript:** Daniel Kondziella, Marwan H. Othman. **Critical review of the manuscript for important intellectual content:** All authors. **Statistical analysis:** Marwan H. Othman, Markus Harboe Olsen. **Obtained funding:** Daniel Kondziella. **Administrative, technical, or material support:** All authors. **Supervision:** Daniel Kondziella, Kirsten Møller, Jesper Kjaergaard. Conflict of Interest Disclosures: The authors report no competing interests. Approval of the manuscript before submission: All authors. **Funding/Support:** This study received funding from Region Hovedstadens Forskningsfond, Rigshospitalets Forskningspuljer, and Offerfonden. The authors are solely responsible for the material, its content, and results. The views expressed in the material are the authors’ own and do not necessarily reflect those of the funders. **Role of the Funder/Sponsor:** The funding sources had no role in the design and conduct of the study; collection, management, analysis, and interpretation of the data; preparation, review, or approval of the manuscript; and decision to submit the manuscript for publication.

## Abstract

**Background:** Identifying residual consciousness in patients with disorders of consciousness (DoC) in the intensive care unit (ICU) is crucial for treatment decisions, but sensitive low-cost bedside markers are missing. We investigated whether automated pupillometry combined with passive and active cognitive paradigms can identify residual consciousness in ICU patients with traumatic or non-traumatic DoC.

**Methods:** In a prospective observational cohort study, clinically low- or unresponsive ICU patients with traumatic and non-traumatic DoC were enrolled from neurological and non-neurological ICUs at a tertiary referral center (Rigshospitalet, Copenhagen University Hospital, Copenhagen, Denmark). Age- and sex-matched healthy volunteers served as controls. Participants with eye disorders were excluded. Patients were categorized into those without (coma or unresponsive wakefulness syndrome, ≤UWS) or with (minimally conscious state or better, ≥MCS) clinical signs of residual consciousness. Using automated pupillometry, we recorded pupillary dilation as a response to passive (visual and auditory stimuli) and active (mental arithmetic) cognitive paradigms, with success criteria depending on the specific task (e.g., ≥3 of 5 pupillary dilations on 5 consecutive mental arithmetic tasks).

**Results:** We obtained 699 pupillometry recordings at 178 time points from 91 brain-injured ICU patients (mean age 60±13.8 years; 31% women; 49.5% non-traumatic brain injuries). Recordings were also obtained from 26 matched controls (59±14.8 years, 38% women). Passive paradigms yielded limited distinctions between patient groups and controls. However, active paradigms involving mental arithmetic enabled discrimination between different states of consciousness. With mental arithmetic of moderate complexity, ≥3 pupillary dilations were seen in 50.0% ≥MCS patients and 17.8% ≤UWS patients (OR 4.56; 95% CI 2.09-10.10, *p*<0.001). In comparison, 76.9% healthy controls responded with ≥3 pupillary dilations (*p*=0.028). Results remained consistent across sensitivity analyses using different thresholds for success. Spearman’s Rank analysis underscored the robust association between pupillary dilations during mental arithmetic and consciousness levels (rho=1, *p*=0.017). Notably, one behaviorally unresponsive patient demonstrated persistent command-following behavior two weeks before overt signs of awareness, suggesting a state of prolonged CMD.

**Conclusions:** Automated pupillometry combined with mental arithmetic can identify cognitive efforts, and hence residual consciousness, in ICU patients with acute DoC.

## Introduction

Identifying residual consciousness in patients with disorders of consciousness (DoC) in the intensive care unit (ICU) is crucial for treatment decisions,^1^ but low-cost sensitive bedside markers are missing.

Automated pupillometry can detect cognitive load by demonstrating pupillary dilations in subjects exposed to arousal or mental activity.^2,3^ For example, in awake volunteers, pupillary dilation occurs each time they engage in mental arithmetic, followed by pupillary constriction when told to relax. If this cycle is successfully repeated, say, five times, an individual has demonstrated command-following abilities without the need for verbal or skeletal muscle motor output.

We hypothesized that automated pupillometry identifies residual consciousness, and potentially cognitive motor dissociation (CMD),^1^ in a subset of ICU patients with acute brain injury and DoC.

## Methods

### Study participants

We prospectively enrolled low-responsive ICU patients with acute traumatic and non-traumatic brain injuries admitted to the neurocritical and cardiological ICUs of a tertiary referral center. The ICUs were screened daily for patients admitted overnight, and patients were consecutively enrolled when deemed feasible by the attending clinicians. Using automated pupillometry, we investigated patients once or, if possible, longitudinally until they recovered consciousness, were discharged from the ICU, or died, whatever came first. Age- and sex-matched healthy volunteers were recruited in parallel by local advertisement. Participants with eye disease of any kind were excluded.

### Clinical classification of consciousness levels

Applying previously described criteria,^4^ DoC patients were classified into coma, unresponsive wakefulness syndrome (UWS), minimally conscious states (MCS) minus/plus, and emerged from MCS at each pupillary measurement. Intravenous sedation was stopped or decreased as much as possible.^4^ Patients were divided into those without (≤UWS) and with (≥MCS) clinical signs of residual consciousness. Age- and sex-matched healthy volunteers were recruited by local advertisement. Subjects with eye disease were excluded.

### Evaluation of pupillary responses to cognitive loads

Utilizing a PLR®-3000 pupillometer (NeurOptics, Laguna Hills, CA, USA; sampling rate 30 Hz, accuracy ± 0.03 mm), we measured pupillary responses over 10 minutes (detailed video in *Supplementary Material*).

#### Passive cognitive paradigms

The first stimulus was the participants’ own facial reflection in a mirror; the second a series of three different 10 seconds long sound clips, each with 20 seconds of white noise interspersed. The first sound clip was Aaron Copland’s Rodeo – Four Dance Episodes,^5^ the second that of a crying toddler, and the third a burglary alarm, all included for their arousal-inducing properties. We used different thresholds to classify pupil dilation as successful. For the mirror stimulus, a single pupillary dilation was sufficient. For the series of pre-recorded sounds, ≥2 of three pupillary dilations were required for success. We compared the pupillary size during the 10 seconds of the stimulus to the 5 seconds immediately before it and to 5 seconds during the mid-resting period (so that the pupil could return to baseline after a stimulus).

#### Active cognitive paradigms

These included two series of mental arithmetic tasks of increasing complexity. We previously showed that such tasks can induce sufficient cognitive load in healthy volunteers and neurological patients detectable by automated pupillometry.^2^ The mental arithmetic tasks consisted of five sets of moderate and five of high complexity (4×36; 8×32; 3×67; 6×37; 7×43, and 21×22; 33×32; 55×54; 43×44; 81×82, respectively). Participants were asked to engage in mental arithmetic and relax, five times in succession. Task duration was set to 20 seconds with 20 seconds rest in-between. Minimal cut-offs for successful command-following were defined as ≥3 pupillary dilations on five mental arithmetic tasks in one set (moderate or hard mental arithmetic). To test more conservative thresholds, we also included analyses using cut-offs of ≥3 pupillary dilations in both sets, and cut-offs of ≥4 pupillary dilations in one and in both sets.

### Pupillary dilation

This was defined as a significantly larger pupil compared to the resting period before and after a stimulus/task. Trigger markers of the applied stimuli were manually inserted while recording. All pupil measurements were included for each period of interest and compared using the Student’s t-test. Although these were autocorrelated measurements, we used the unpaired t-test because it is more conservative and does not require equal numbers of measurements in the groups. All recordings were visually inspected for artifacts; however, presence or absence of pupillary dilations were revealed to the investigators only after the publication of the statistical analysis plan.^6^ Data points with abrupt deviations exceeding 1.5 mm from the preceding data point (i.e., blinks) were labelled as physiologically implausible and removed. For transparency, we have added a supplemental file revealing all corrected trigger markers (Supplemental material).

### Sample size and power calculations

The study sample size was calculated based on data obtained from a feasibility study.^2^ To detect a clinically significant difference in pupillary dilations between resting and stimulation periods, we aimed for a statistical power of 80% and a type 1 error probability (α) of 0.05. This calculation resulted in a minimum required sample size of n=41 patients. Because the pupillometry paradigm was easy to implement in daily clinical routine and associated with nor harm or risks and because we also wanted to test for more conservative success thresholds, we aimed for a sample size approximately twice that number.

### Statistics analysis

Following a preregistered statistical analysis plan,^6^ statistics were performed in R (R Core Team, Vienna, Austria). The number of pupillary dilations of each group were compared using the chi-squared test or Fisher’s exact test as appropriate. Numeric data were analyzed for intergroup comparisons using Student’s t-test, Mann–Whitney U-test, or Kruskal–Wallis test. Spearman’s Rank analysis was done to assess the association between pupillary dilations count during mental arithmetic and consciousness levels. *P*≤0.05 was considered significant.

### Data sharing statement and code availability

Study data are available in the article and Supplementary material (raw data will be made available upon final journal publication of the paper). The algorithm for pupillometry data processing is available at https://github.com/lilleoel/clintools.

### Additional material

Details are provided (i) in a preregistered statistical analysis plan at Zenodo.org,^6^ (ii) in the online *Supplementary Material*, including pupillometry raw data, anonymized patient data, and a videoclip of the examination setting, and (iii) at https://cran.r-project.org/web/packages/clintools/, which also includes the code for processing the pupillometry data. We also provide a step-by-step guide to use automated pupillometry for the bedside detection of covert consciousness (*Supplementary Methods*).

### Ethics

Regional Ethics Committee (Region Hovedstaden) approval was obtained (H-21022096).

## Results

We obtained 699 pupillometry recordings at 178 time points from 91 brain-injured ICU patients (mean age 60±13.8 years; 31% women; 49.5% non-traumatic brain injuries); 95.5% were ≤UWS at first evaluation. Fifty-four participants were investigated twice, and 28 three times. Recordings were also obtained from 26 age- and sex-matched controls (59±14.8 years, 38% women). **Table 1** and **Tables S1-S4** provide baseline statistics, individual patient data, and detailed pupillometry results.

**Table 1.**
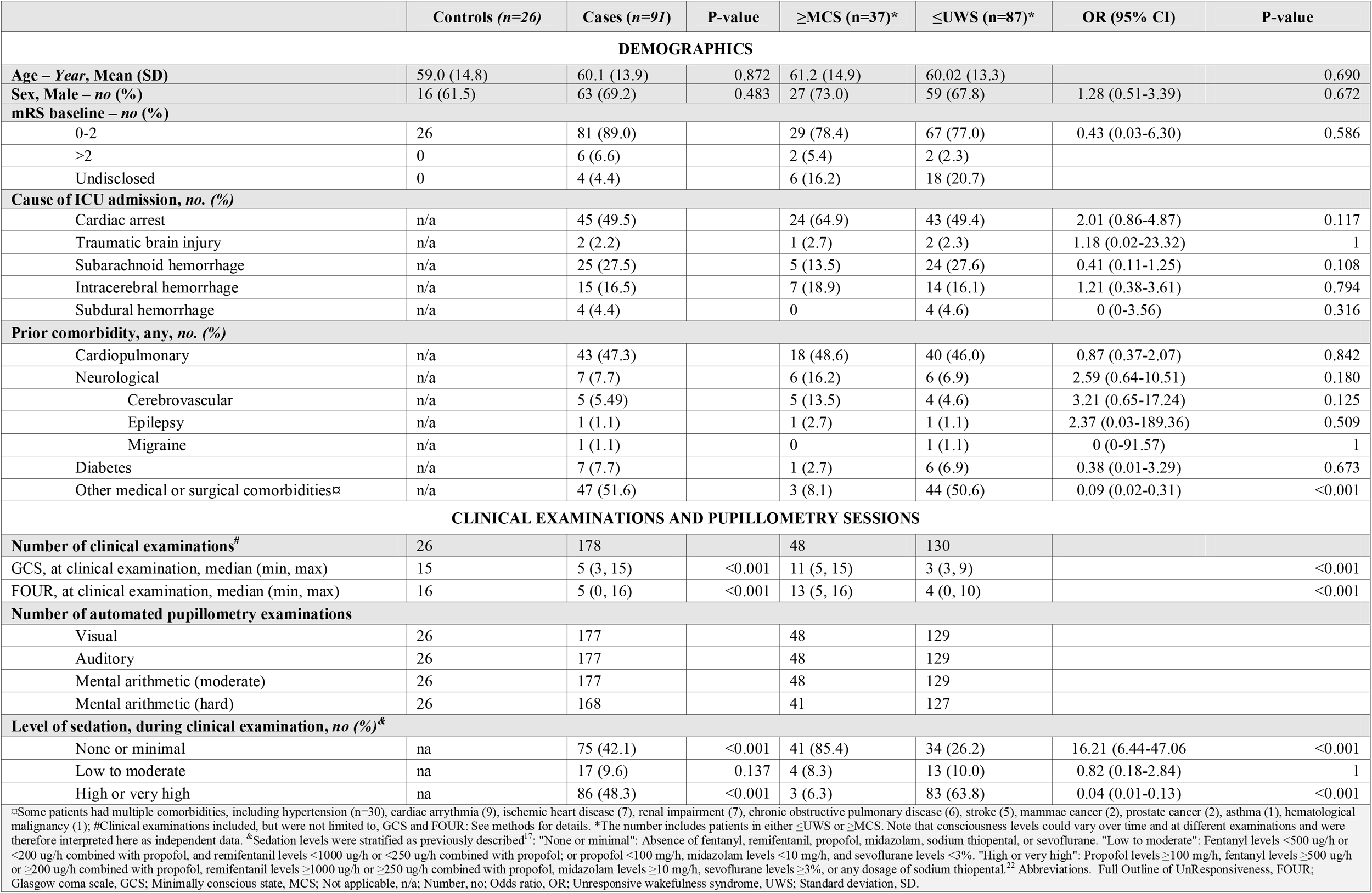
Baseline Characteristics of Study Population.

### Passive cognitive paradigms

Pupillary dilations in the mirror paradigms differed between healthy controls and patients (**Figure 1)**. Controls responded to their reflection more often than ≥MCS patients (odds ratio (OR) 4.93; 95% CI 1.55-16.69, *p*=0.003). However, there was no difference between ≤UWS and ≥MCS patients. Auditory stimuli were not different between healthy controls and patients, and neither between ≤UWS and ≥MCS groups.

**Figure 1.**
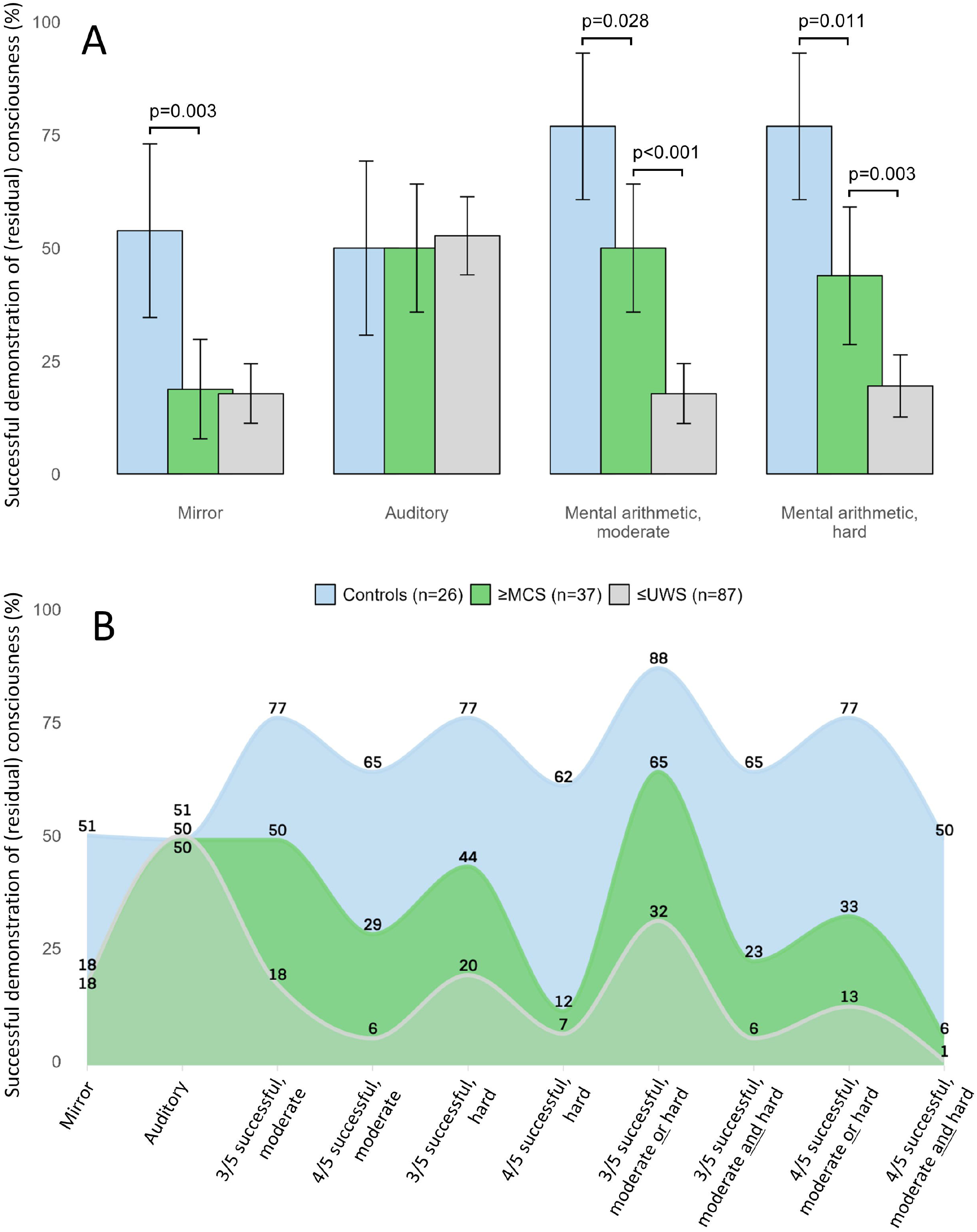
Pupillary responses to passive and active paradigms in controls, ≥MCS and ≤UWS patients. **(A)**. The bar chart illustrates the pupillary responses to different automated pupillometry paradigms, both passive (visual, auditory) and active (mental arithmetic), along with the corresponding number of patients and controls (%) who achieved success in each paradigm. **Area chart of pupillary responses (%) to passive paradigms and active task-based paradigms with adjusted thresholds for success (B)**. The area chart depicts the proportions (%) of patients with acute brain injury and controls achieving detectable pupillary dilations in response to passive visual and auditory paradigms, and active mental arithmetic paradigms. The latter included five sets of moderate and five of high complexity mental arithmetic (moderate: 4×36; 8×32; 3×67; 6×37; 7×43; hard: 21×22; 33×32; 55×54; 43×44; 81×82). eMCS, emerged from MCS; MCS, minimally conscious state; UWS, unresponsive wakefulness syndrome

### Active cognitive paradigms

Pupillometry with mental arithmetic could distinguish between patient groups (**Figure 1**). With mental arithmetic of moderate complexity, ≥3 pupillary dilations were seen in 50.0% ≥MCS patients and 17.8% ≤UWS patients (OR 4.56; 95% CI 2.09-10.10, *p*<0.001). In comparison, 76.9% healthy volunteers responded with ≥3 pupillary dilations during mental arithmetic, different from patients (*p*=0.028). With increasingly complex mental arithmetic, the ability of pupillometry to distinguish between healthy controls, ≤UWS patients and ≥MCS patients remained unchanged. Spearman’s Rank analysis underscored the association between pupillary dilations during mental arithmetic and consciousness levels (rho=1, *p*=0.017; **Figure 2**). These differences remained consistent with a more conservative cut-off of ≥4 pupillary dilations per five mental arithmetic tasks in *either* one of the hard or the moderate complex set of arithmetic tasks, and in *both* hard and moderate complex set of tasks. However, with a cut-off of ≥4 pupillary dilations, the percentage of success in ≤UWS patients decreased to 6.2% and 7.1% for moderate- and hard-level arithmetic tasks, respectively. The strictest criteria for success required ≥4 pupillary dilations in both sets of arithmetic tasks; only one patient (0.8%) achieved this. **Figure 3A** shows representative examples.

**Figure 2.**
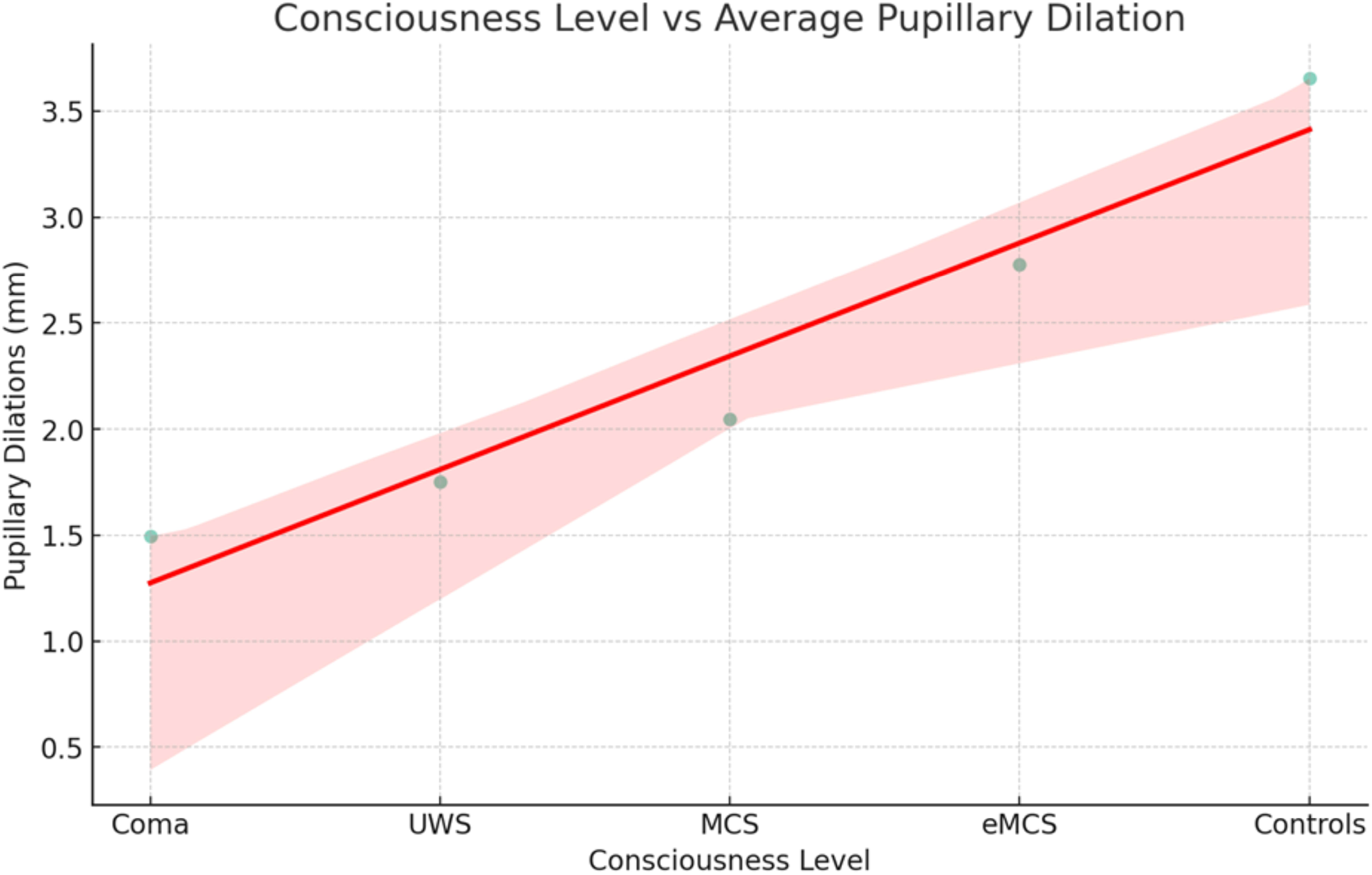
Scatter Plot of the correlation between consciousness levels and average pupillary dilations during mental arithmetic. This graph visualizes the relationship between the average number of pupillary dilations during mental arithmetic in millimeters (calculated as the mean of medium- and hard-task pupillary dilations) and coded consciousness levels in patients. The red line represents a linear regression fit, indicating the trend of correlation.

**Figure 3.**
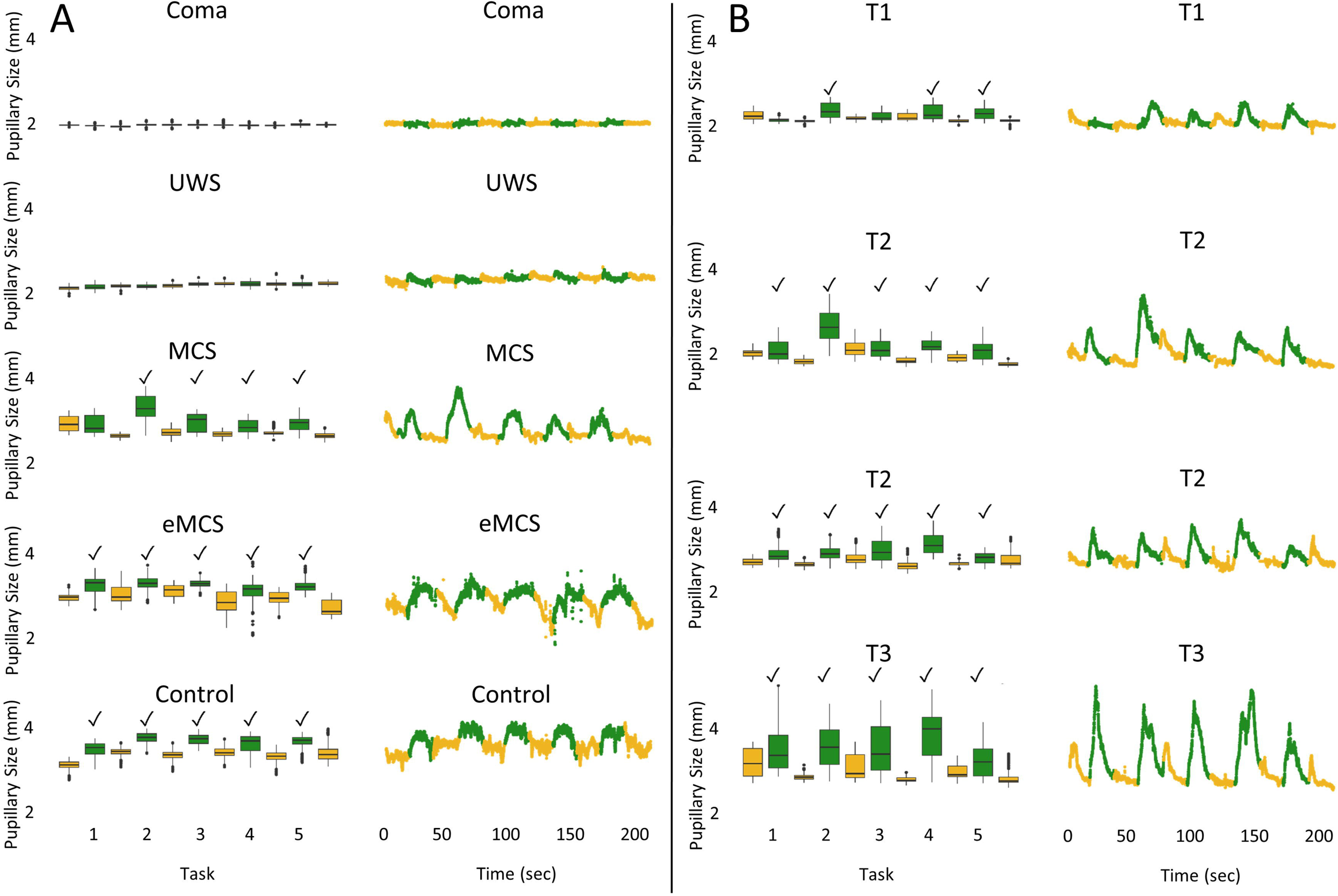
Pupillometry data from four patients in different states of consciousness and one control (A), as well as one patient with covert consciousness (B). This figure illustrates pupillometry data obtained from participants in various states of consciousness. The data reflects the count of successful pupillary dilations recorded through automated pupillometry during mental arithmetic paradigms. The figure presents data for each subject in two distinct formats: boxplots (left) and scatter plots (right). The color code distinguishes between periods of mental arithmetic (green) and rest periods (yellow), with the x-axis representing time in seconds (“0-50-100-150-200”). The data indicate that pupillary sizes during mental arithmetic were significantly larger than during rest periods, indicating consistent pupillary dilation. This was observed in at least four out of the five tasks for participants in MCS, eMCS, and the healthy volunteer, as well as a patient in presumed CMD (whose case description is given in Figure 3). However, there were no significant differences between task and rest periods for the coma and UWS patients. “✓” denotes significant pupillary dilation (p-value < 0.0001); mm, millimeter.

### Stratification by injury type

When stratified by injury type, differences persisted between patients with anoxic brain damage with both moderate and complex mental arithmetic. With non-anoxic brain injuries, the differences were numerically, but not statistically, different (**Table S5**).

### Prolonged CMD

In one behaviorally UWS patient with subarachnoid hemorrhage, repeated pupillary dilations during mental arithmetic were detected two weeks before overt command following noted by the attending clinicians, suggesting prolonged CMD (**Figure 3B, Figure 4**).

**Figure 4.**
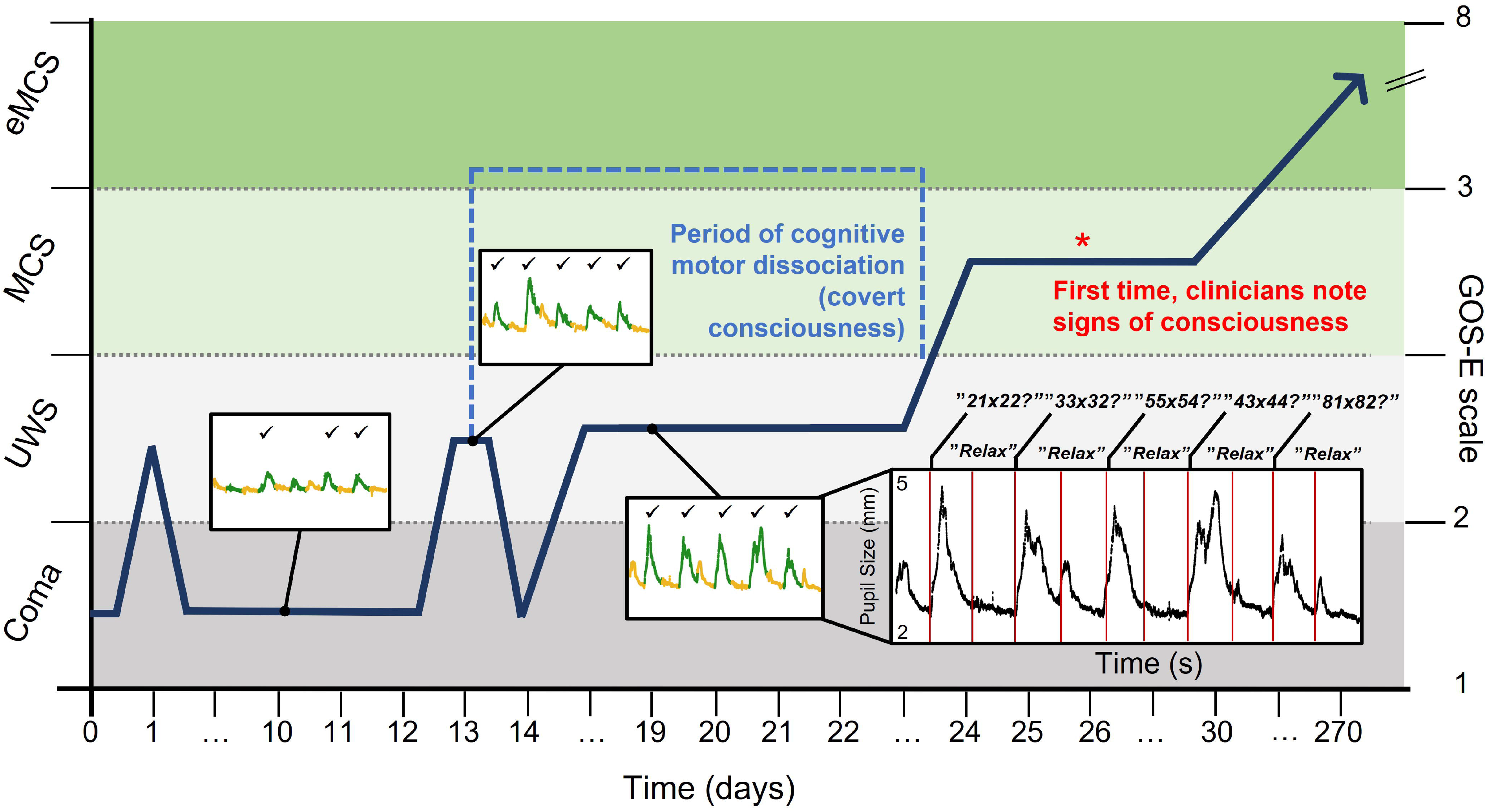
Covert consciousness detected by automated pupillometry 2 weeks prior to overt command-following. A previously healthy woman (age range 36-40 years) with subarachnoid hemorrhage was treated with clipping of the responsible pericallosal artery aneurysm. The diagram depicts the trajectory of consciousness recovery (dark blue arrow), including mental arithmetic-task based pupillometry on days 10, 13 and 19. Between days 0-24, the patient’s clinical arousal level (y-axis) indicated she was in a coma or UWS. On day 10, mental arithmetic performance (3 of 5 pupillary dilations) reached the prespecified threshold for command following, indicative of CMD. The patient successfully followed commands also on days 13 and 19, as shown by 5/5 pupillary dilations. This period of covert consciousness lasted for almost 2 weeks (light blue dashed line). First on day 26 did the treating clinicians notice visual tracking and attempts to follow commands (red asterix). At 9-month follow-up, the patient could attend most activities of daily living (Glasgow Outcome Scale-Extended score of 5). Inserts: Green lines of the plots represents the stimulation periods, while the yellow lines represent the rest periods in between tasks; check marks in the plots indicate presence of pupillary dilations during mental arithmetic tasks; y-axis depicts pupillary diameter in millimeter, x-axis time in seconds. See also Figure 3B. eMCS, emerged from MCS; MCS, minimally conscious state; UWS, unresponsive wakefulness syndrome.

## Discussion

This is, to our knowledge, the first study investigating automated pupillometry combined with passive and active cognitive paradigms to discover residual consciousness in acute DoC. In contrast to fMRI- and EEG-based consciousness paradigms which are computationally complex, labor-intensive, and logistically demanding (precluding their routine use in the ICU^6^), pupillometry only requires a low-cost, handheld bedside device and a relatively simple data processing algorithm. Our key finding is that pupillometry together with mental arithmetic could detect residual consciousness in acute DoC, including persistent command-following behavior several days prior to overt awareness.

### Passive cognitive paradigms cannot distinguish between degrees of DoC

While seeing one’s own facial reflection in a mirror caused pupillary dilation more than twice as often in conscious volunteers than in patients, this paradigm could not distinguish between patient groups. The auditory paradigm worked even worse; there were no differences between controls and patients. The most reasonable explanation is that pupillary dilation requires sustained arousal or prolonged mental activity that exceeds that provoked by passive visual and auditory stimuli. Although this may appear disappointing at first sight, there is also a very reassuring aspect to this negative finding, as explained in the next section on active paradigms.

### Active cognitive paradigms can differentiate between degrees of DoC

Pupillometry combined with mental arithmetic revealed strong differences in the response to cognitive loads, from absence of responses in most comatose and UWS patients to presence of responses in most conscious volunteers, with MCS patients ranging in between. Results remained consistent across sensitivity analyses using different thresholds for success. Reassuringly, given that passive paradigms did not result in meaningful group differences, we can confidently rule out that the pupillary responses seen with mental arithmetic were induced by passively hearing the instructions. Instead, they do seem to reflect true cognitive load and hence, mental activity. Remarkably, in at least one behaviorally UWS patient such mental activity was observed on three occasions almost two weeks before clinical signs of consciousness were noted by the attending clinicians, demonstrating the proof-of-concept that automated pupillometry paradigm can detect states suggestive of CMD.

The success rate of 62-88% in healthy volunteers is identical to that seen in active cognitive paradigms utilizing fMRI^7^ and EEG.^8^ The low success rate contributes to decreased sensitivity, a limitation that also applies to fMRI- and EEG-based paradigms. The optimal threshold for success in terms of sensitivity and specificity remain unknown. There is no gold standard to measure consciousnees,^9^ but CMD occurs in 15-20% of acute^10,11^ and chronic^12^ DoC patients, suggesting that ≥4 dilations in the moderate or the hard mental arithmetic task may be the best threshold.

Although our method is unlikely to have identified every patient with residual or covert consciousness, we think it potentially provides clinically meaningful positive predictive values and specificities. A caveat is that the positive results mainly came from patients with anoxic brain injury. Whether pupillometry with mental arithmetic works equally well in acute DoC caused by traumatic and other non-anoxic brain injuries remains to be seen. Notably, however, pupillometry with mental arithmetic appears to have greater potential than pupillometry combined with the auditory ‘local global’ paradigm,^13^ probably because of the larger cognitive load that comes with mental arithmetic. Head-to-head studies comparing automated pupillometry with fMRI- and EEG-based active paradigms are required to investigate this further. In the meantime, we provide detailed advice on how to replicate our methodology, including a preregistered statistical analysis plan, video instructions regarding the examination technique (upon final journal publication), and open access to the code and algorithm required to process the pupillometry data.

## Conclusions

Automated pupillometry combined with mental arithmetic can discover residual consciousness in acute DoC patients, potentially including those with CMD. Given that fMRI- and EEG-based paradigms to identify residual consciousness depend on sophisticated computational expertise and are logistically challenging in the ICU, we think the convenience of this low-cost bedside paradigm makes it a promising novel biomarker in acute DOC.

## Supporting information

STROBE guidelines

## Data Availability

All study data are available in the article and Supplementary material (including raw data). The algorithm for pupillometry data processing is available at https://github.com/lilleoel/clintools.

## References

1. Claassen J, Kondziella D, Alkhachroum A, et al. Cognitive Motor Dissociation: Gap Analysis and Future Directions. Neurocrit Care. Published online 2023. doi:10.1007/S12028-023-01769-3

2. Vassilieva A, Olsen MH, Peinkhofer C, Knudsen GM, Kondziella D. Automated pupillometry to detect command following in neurological patients: A proof-of-concept study. PeerJ. 2019;7:e6929(5):Doi.org/10.7717/peerj.6929. doi:10.7717/peerj.6929

3. Stoll J, Chatelle C, Carter O, Koch C, Laureys S, Einhäuser W. Pupil responses allow communication in locked-in syndrome patients. Curr Biol. 2013;23(15):R647–R648. doi:10.1016/j.cub.2013.06.011

4. Amiri M, Fisher PM, Raimondo F, et al. Multimodal prediction of residual consciousness in the intensive care unit: the CONNECT-ME study. Brain. 2023;146(1):50–64. doi:10.1093/BRAIN/AWAC335

5. Naci L, Owen AM. Making every word count for nonresponsive patients. JAMA Neurol. 2013;70(10):1235–1241. doi:10.1001/jamaneurol.2013.3686

6. Othman MH, Olsen MH, Møller K, Kjaergaard J, Kondziella D. Automated pupillometry to uncover signs of consciousness in acute brain injury: statistical analysis plan. Published 2023. https://zenodo.org/records/6627565

7. Monti MM, Vanhaudenhuyse A, Coleman MR, et al. Willful Modulation of Brain Activity in Disorders of Consciousness. N Engl J Med. 2010;362(7):579–589. doi:10.1056/NEJMoa0905370

8. Cruse D, Chennu S, Chatelle C, et al. Bedside detection of awareness in the vegetative state: a cohort study. Lancet. 2011;378(9809):2088–2094. doi:10.1016/S0140-6736(11)61224-5

9. Kondziella D, Bender A, Diserens K, et al. European Academy of Neurology guideline on the diagnosis of coma and other disorders of consciousness. Eur J Neurol. 2020;27(5):741–756. doi:10.1111/ene.14151

10. Claassen J, Doyle K, Matory A, et al. Detection of Brain Activation in Unresponsive Patients with Acute Brain Injury. N Engl J Med. 2019;380(26):2497–2505. doi:10.1056/NEJMoa1812757

11. Egbebike J, Shen Q, Doyle K, et al. Cognitive-motor dissociation and time to functional recovery in patients with acute brain injury in the USA: a prospective observational cohort study. Lancet Neurol. 2022;21(8):704–713. doi:10.1016/S1474-4422(22)00212-5

12. Kondziella D, Friberg CK, Frokjaer VG, Fabricius M, Møller K. Preserved consciousness in vegetative and minimal conscious states: systematic review and meta-analysis. J Neurol Neurosurg Psychiatry. 2016;87(5):485–492. doi:10.1136/jnnp-2015-310958

13. Sangare A, Quirins M, Marois C, et al. Pupil dilation response elicited by violations of auditory regularities is a promising but challenging approach to probe consciousness at the bedside. Sci Rep. 2023;13(1). doi:10.1038/S41598-023-47806-1

