## Supplementary Material and Methods for "Automated pupillometry to detect residual consciousness in acute brain injury"

CONTENT

1. **Supplemental Methods I:** Practical guide to automated pupillometry
2. **Supplemental Table 1:** Individualized Demographical Data
3. **Supplemental Table 2:** Individualized Clinical Data
4. **Supplemental Table 3:** Individualized Pupillometry Results
5. **Supplemental Table 4:** Average Pupillary Dilation (mm) in response to passive and active paradigms across different consciousness levels.
6. **Supplemental Table 5:** Stratified group comparisons - successful pupillary dilations in response to passive and active paradigms.

**Supplemental Methods I**

### *A step-by-step guide to use automated pupillometry for the bedside detection of covert consciousness*

Automated pupillometry (PLR®-3000 Pupillometer) is a simple and handy device used for measuring pupillary size longitudinally. When paired with active paradigms, we found it to be useful in detecting covert consciousness in ICU DoC patients. The following is a guide on how to employ this method in practice.

1. **Device settings and preparations**

Power on the device and set the protocol to “Extended Mode (Max Duration 10 min)”. Enter the patient's ID, along with the current time and date. This step aids in distinguishing between patients when multiple recordings are gathered, ensuring precise data management.

If pre-recorded instructions are not available, it's important to have a timer on hand to precisely time when to deliver your verbal instructions. Allocate 20 seconds for each arithmetic task, and follow the pattern depicted in the figure below, which also includes 20-second resting intervals between tasks.

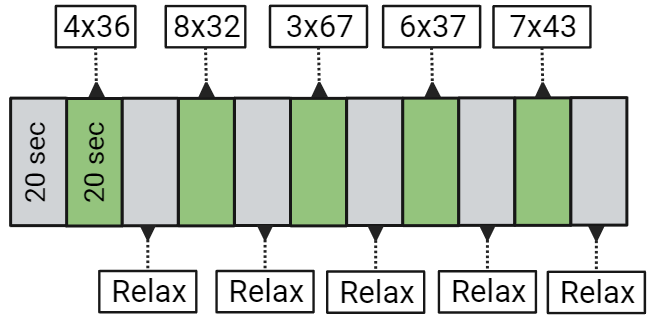

**Figure 1: Mental arithmetic paradigm with moderate-level tasks**

It is essential to provide the patient with clear instructions regarding the paradigm. Inform the patient that you will gently open their eye and measure the pupillary size while presenting them with a series of five arithmetic tasks. The primary goal is to assess their level of awareness. Stress that this procedure is entirely painless and will not cause any discomfort.

Be sure to explicitly ask the patient to concentrate and perform to the best of their abilities during each arithmetic task. Additionally, advise the patient to relax when prompted during the intervals between tasks.

2. **Data recording**

Position yourself appropriately and, if feasible, orient the patient's head towards the examiner. A helpful tip: consider placing a pillow beneath your arms to alleviate strain on shoulders.

[image removed to comply with MedRxiv instructions; material available on request from the authors]

**Picture 1: Bedside arrangement (simulation)**

Now, align the pupillometer with either the right or left eye as convenient. Start the recording process by pressing and holding the corresponding button (e.g., "right" button for the right eye) until the pupil is captured, indicated by a green or yellow circle. Release your finger from the button to initiate pupil recording.

For each 20-second interval, provide a verbal instruction and use the "arrow up" or "arrow down" button to insert a marker. Once the paradigm is completed, tap the "right" button to conclude the recording.

3. **Data processing**

Retrieve the data in the form of an XLS file using the device's Bluetooth function. Please note that this device is only compatible with Windows systems.

Upload the XLS file to R Studio and execute the script provided in the Supplementary Material. Make sure to install and load the 'clintools' package from <https://cran.r-project.org/web/packages/clintools/> to ensure the script functions correctly.

Assuming that the distribution of triggers and commands aligns with the described methodology, these scripts should generate scatterplots illustrating the frequency of pupillary dilations.

For questions, please feel free to contact the authors.

| **Supplemental Table 1: Individualized Demographical Data** | | | | | | | |
| --- | --- | --- | --- | --- | --- | --- | --- |
| **ID** | **Etiology** | **Age*** | **Sex** | **mRS** | **History of Comorbidities** | **Discharge**  **Consciousness level** | **Discharge**  **Survival status** |
| 1 | SAH | X | Male | 1 |  | Coma | Alive |
| 2 | ICH | X | Male | 1 | HTN, Cardiac Arrythmia | LIS | Alive |
| 3 | ICH | X | Female | 1 | CRD | eMCS | Alive |
| 4 | SAH | X | Female | 1 | Renal impairment | MCS+ | Alive |
| 5 | OHCA | X | Male | 1 | HTN, Prostate cancer | FC | Alive |
| 6 | OHCA | X | Male | 1 |  | FC | Alive |
| 7 | SDH | X | Male | 1 | T2D, Cardiac Arrythmia, Hematologic cancer | Coma | Alive |
| 8 | OHCA | X | Female | 1 | HTN, HLD, CRD | FC | Alive |
| 9 | OHCA | X | Male | 1 | HTN, Renal impairment | eMCS | Deceased |
| 10 | SAH | X | Female | 1 | HTN | MCS+ | Alive |
| 11 | OHCA | X | Male | 1 | Epilepsy | FC | Alive |
| 12 | ICH | X | Male | 1 | Stroke | MCS+ | Alive |
| 13 | OHCA | X | Male | 1 |  | FC | Alive |
| 14 | OHCA | X | Female | 1 | Cardiac Arrythmia | FC | Alive |
| 15 | OHCA | X | Male | 1 |  | FC | Alive |
| 16 | SAH | X | Male | 3 | HTN, HLD, Cardiac Arrythmia, Stroke | eMCS | Alive |
| 17 | ICH | X | Male | 1 | HTN, Renal impairment | Coma | Deceased |
| 18 | OHCA | X | Male | 1 |  | FC | Alive |
| 19 | SAH | X | Male | 1 |  | CS | Alive |
| 20 | SAH | X | Female | 1 |  | MCS+ | Alive |
| 21 | OHCA | X | Male | 1 | HTN, IHD/AMI | UWS | Deceased |
| 22 | SAH | X | Female | 1 |  | eMCS | Alive |
| 23 | SAH | X | Female | 1 |  | MCS- | Alive |
| 24 | OHCA | X | Male | 1 | Cardiac Arrythmia | CS | Alive |
| 25 | OHCA | X | Male | 1 | HTN | FC | Alive |
| 26 | SAH | X | Male | 1 |  | CS | Alive |
| 27 | SAH | X | Male | 3 |  | UWS | Deceased |
| 28 | OHCA | X | Female | 1 | Breast cancer | FC | Alive |
| 29 | OHCA | X | Female | 1 | HLD | FC | Alive |
| 30 | SAH | X | Male | 1 |  | CS | Alive |
| 31 | OHCA | X | Male | 1 |  | CS | Alive |
| 32 | OHCA | X | Male | 1 |  | FC | Alive |
| 33 | OHCA | X | Male | 1 |  | UWS | Deceased |
| 34 | OHCA | X | Male | 1 | HLD | UWS | Deceased |
| 35 | ICH | X | Male | 1 | HTN, CRD, Stroke | eMCS | Alive |
| 36 | SAH | X | Female | 1 | HTN, | UWS | Deceased |
| 37 | SDH | X | Male | 1 |  | MCS+ | Alive |
| 38 | SAH | X | Female | 1 |  | Coma | Alive |
| 39 | OHCA | X | Male | 2 |  | FC | Alive |
| 40 | OHCA | X | Female | 1 | IHD/AMI, Renal impairment, Breast cancer | eMCS | Alive |
| 41 | OHCA | X | Male | 1 |  | FC | Alive |
| 42 | OHCA | X | Male | 1 |  | eMCS | Alive |
| 43 | OHCA | x | Male | 7 | HTN, HLD, IHD/AMI | Coma | Deceased |
| 44 | OHCA | X | Male | 1 | HTN, HLD, IHD, CRD, Renal impairment | CS | Alive |
| 45 | OHCA | X | Female | 7 |  | Coma | Deceased |
| 46 | SAH | X | Female | 1 | HTN | UWS | Alive |
| 47 | OHCA | X | Male | 1 |  | Coma | Deceased |
| 48 | TBI | X | Male | 1 |  | UWS | Alive |
| 49 | ICH | X | Male | 1 | Stroke | MCS- | Alive |
| 50 | OHCA | X | Male | 1 | HTN, HLD | FC | Alive |
| 51 | SAH | X | Female | 3 | HTN, Cardiac Arrythmia | MCS- | Alive |
| 52 | OHCA | X | Male | 7 |  | CS | Deceased |
| 53 | SAH | X | Female | 1 | HTN, HLD | UWS | Alive |
| 54 | SAH | X | Female | 1 |  | Coma | Deceased |
| 55 | OHCA | X | Male | 4 |  | eMCS | Alive |
| 56 | OHCA | X | Female | 1 | HTN, | eMCS | Alive |
| 57 | SAH | X | Female | 1 | Migraine | Coma | Deceased |
| 58 | SAH | X | Female | 1 | HTN, | MCS- | Alive |
| 59 | OHCA | X | Male | 1 | HTN, IHD/AMI, Renal impairment | UWS | Alive |
| 60 | OHCA | X | Male | 1 |  | FC | Alive |
| 61 | ICH | X | Male | 1 |  | UWS | Alive |
| 62 | SAH | X | Male | 1 |  | UWS | Alive |
| 63 | ICH | X | Male | 1 | IHD/AMI | MCS+ | Alive |
| 64 | ICH | X | Male | 1 | Prostate cancer | MCS+ | Alive |
| 65 | TBI | X | Male | 1 |  | MCS- | Alive |
| 66 | ICH | X | Male | 3 |  | Coma | Alive |
| 67 | OHCA | X | Male | 7 |  | UWS | Deceased |
| 68 | OHCA | X | Male | 4 |  | CS | Alive |
| 69 | SDH | X | Female | 1 |  | Coma | Alive |
| 70 | SAH | X | Male | 1 | HTN, | UWS | Alive |
| 71 | SDH | X | Male | 1 | IHD/AMI | MCS- | Alive |
| 72 | SAH | X | Female | 1 | HTN, | Coma | Deceased |
| 73 | OHCA | X | Male | 1 | HTN, | FC | Alive |
| 74 | OHCA | X | Male | 2 |  | UWS | Alive |
| 75 | SAH | X | Male | 1 |  | CS | Alive |
| 76 | OHCA | X | Male | 2 |  | FC | Alive |
| 77 | SAH | X | Male | 1 |  | Coma | Deceased |
| 78 | OHCA | X | Male | 2 |  | Coma | Deceased |
| 79 | OHCA | X | Male | 1 |  | FC | Alive |
| 80 | OHCA | X | Male | 1 | HTN, | FC | Alive |
| 81 | OHCA | X | Male | 1 |  | FC | Alive |
| 82 | ICH | X | Female | 2 | HTN, Stroke | MCS+ | Alive |
| 83 | OHCA | X | Female | 1 | Cardiac Arrythmia | UWS | Deceased |
| 84 | ICH | X | Male | 1 | HTN, CRD | Coma | Deceased |
| 85 | ICH | X | Female | 2 |  | Coma | Alive |
| 86 | OHCA | X | Male | 1 | Atrial fibrillation | CS | Alive |
| 87 | ICH | X | Female | 1 | HTN, | Coma | Deceased |
| 88 | OHCA | X | Male | 1 | HTN, Cardiac Arrythmia, CRD | UWS | Alive |
| 89 | OHCA | X | Male | 1 | HLD, Cardiac Arrythmia, CRD | Coma | Deceased |
| 90 | ICH | X | Male | 1 | HTN | Coma | Alive |
| 91 | SAH | X | Female | 1 | HTN, IHD/AMI, CRD | UWS | Alive |
| Abbreviations: AMI=Acute Myocardial Infarction; CRD=Chronic Respiratory Disease (i.e., asthma or chronic obstructive pulmonary disease); CS=Confusional State; eMCS=emerged from MCS; FC=Fully Conscious State; HLD=Hyperlipidemia; HTN=Hypertension; ICH=Intracerebral Hemorrhage; IHD=Ischemic Heart Disease; LIS=Locked-in Syndrome; MCS=Minimally Conscious State (minus “-“/plus “+”); mRS=modified Rankin Scale; OHCA=Out of Hospital Cardiac Arrest; SAH=Subarachnoid bleeding; SDH=Subdural Hemorrhage; TBI=Traumatic Brain Injury; UWS=Unresponsive Wakefulness Syndrome. *Age was removed to comply with MedRxiv instructions. | | | | | | | |

| **Supplemental Table 2: Individualized Clinical Data** | | | | | | | |
| --- | --- | --- | --- | --- | --- | --- | --- |
| **ID** | **Clinical Examination** | **Admission Days** | **Ventilator** | **Sedation Level*** | **FOUR** | **GCS** | **Consciousness Level** |
| 1 | 1 | 9 | tracheostomy | F=300 µg/h; MI=25 mg/h | E0, M0, B2, R0 | 3 | Coma |
|  | 2 | 16 | tracheostomy |  | E0, M2, B2, R1 | E1, V1, M4 | Coma |
| 2 | 1 | 2 | intubated | P=50 mg/h; RE=750 µg/h | E4, M4, B4, R1 | E4, V1, M6 | LIS |
| 3 | 1 | 1 | intubated | P=280 mg/h; RE=700 µg/h | E0, M0, B2, R0 | 3 | Coma |
|  | 2 | 8 | tracheostomy |  | E0, M2, B2, R1 | E1, V1, M4 | Coma |
|  | 3 | 10 | tracheostomy |  | E1, M2, B2, R1 | E2, V1, M4 | UWS |
| 4 | 1 | 2 | intubated | F=500 µg/h; MI=30 mg/h; T=200 mg/h | E0, M0, B2, R0 | 3 | Coma |
|  | 2 | 9 | tracheostomy |  | E2, M4, B4, R1 | E3, V1, M6 | MCS+ |
|  | 3 | 12 | tracheostomy |  | E2, M4, B4, R1 | E3, V1, M6 | MCS+ |
| 5 | 1 | 0 | intubated | F=200 µg/h; P=80 mg/h | E0, M0, B2, R0 | 3 | Coma |
|  | 2 | 2 | none |  | 16 | E4, V4, M6 | CS |
|  | 3 | 3 | none |  | 16 | 15 | FC |
| 6 | 1 | 3 | intubated | RE=1500 µg/h | E2, M3, B4, R0 | E3, V1, M5 | MCS- |
|  | 2 | 7 | none |  | 16 | 15 | FC |
| 7 | 1 | 4 | intubated | RE=150 µg/h | E0, M2, B4, R1 | E1, V1, M4 | Coma |
| 8 | 1 | 1 | intubated | F=250 µg/h; P=100 mg/h | E0, M0, B4, R0 | 3 | Coma |
| 9 | 1 | 0 | intubated | F=100 µg/h; P=40 mg/h | E0, M0, B4, R0 | 3 | Coma |
| 10 | 1 | 15 | intubated | P=200 mg/h; RE=800 µg/h | E0, M2, B4, R0 | E1, V1, M4 | Coma |
| 11 | 1 | 0 | intubated | F=250 µg/h; P=50 mg/h | E0, M0, B2, R0 | 3 | Coma |
|  | 2 | 3 | none |  | 16 | 15 | FC |
| 12 | 1 | 6 | intubated | P=250 mg/h; RE=1250 µg/h | E0, M0, B4, R0 | 3 | Coma |
|  | 2 | 17 | tracheostomy |  | E4, M4, B4, R1 | E3, V1, M6 | MCS+ |
| 13 | 1 | 1 | intubated | F=200 µg/h; P=200 mg/h | E0, M0, B2, R0 | 3 | Coma |
|  | 2 | 4 | none |  | 16 | E4, V4, M6 | CS |
| 14 | 1 | 1 | intubated | F=200 µg/h; P=120 mg/h | E0, M0, B4, R0 | 3 | Coma |
|  | 4 | 5 | none |  | 16 | 15 | FC |
| 15 | 1 | 0 | intubated | P=150 mg/h; F=200 µg/h | E0, M2, B4, R0 | E1, V1, M4 | Coma |
|  | 2 | 4 | none |  | 16 | 15 | FC |
| 16 | 1 | 5 | tracheostomy | RE=500 µg/h | E0, M2, B4, R1 | E1, V1, M4 | MCS- |
| 17 | 1 | 2 | tracheostomy | F=400 µg/h; MI=20 mg/h | E0, M0, B0, R0 | 3 | Coma |
| 18 | 1 | 1 | intubated | P=150 mg/h; RE=500 µg/h | E0, M2, B4, R0 | E1, V1, M4 | Coma |
|  | 2 | 3 | none |  | E4, M4, B4, R0 | 15 | FC |
| 19 | 1 | 6 | intubated | F=300 µg/h; MI=30 mg/h | E0, M0, B4, R0 | 3 | Coma |
| 20 | 1 | 10 | intubated | F=300 µg/h; MI=30 mg/h | E0, M0, B2, R0 | 3 | Coma |
|  | 2 | 13 | intubated | F=400 µg/h; MI=30 mg/h | E2, M0, B4, R0 | E3, V1, M1 | UWS |
|  | 3 | 19 | tracheostomy |  | E1, M2, B4, R1 | E2, V1, M4 | UWS |
|  | 4 | 24 | tracheostomy |  | E1, M2, B4, R1 | E2, V1, M4 | UWS |
| 21 | 1 | 1 | intubated | F=100 µg/h; P=200 mg/h | E0, M0, B4, R0 | 3 | Coma |
| 22 | 1 | 3 | intubated | F=300 µg/h; MI=30 mg/h | E0, M0, B4, R0 | 3 | Coma |
|  | 2 | 15 | tracheostomy |  | E4, M4, B4, R1 | E3, V1, M6 | MCS+ |
| 23 | 1 | 12 | intubated | F=200 µg/h; MI=12.5 mg/h | E0, M0, B4, R1 | 3 | Coma |
|  | 2 | 16 | tracheostomy |  | E1, M2, B2, R1 | E2, V1, M4 | UWS |
|  | 3 | 19 | tracheostomy |  | E1, M2, B3, R1 | E2, V1, M4 | UWS |
| 24 | 1 | 1 | intubated | F=300 µg/h; P=250 mg/h | E0, M0, B4, R0 | 3 | Coma |
|  | 2 | 4 | none |  | 16 | E4, V4, M6 | CS |
| 25 | 1 | 0 | intubated | F=200 µg/h; P=200 mg/h | E0, M0, B4, R0 | 3 | Coma |
|  | 3 | 4 | none |  | 16 | E4, V4, M6 | CS |
| 26 | 1 | 5 | intubated | P=300 mg/h; RE=2000 µg/h | E0, M2, B4, R0 | E0, V1, M4 | Coma |
| 27 | 1 | 3 | intubated | RE=1000 µg/h | E0, M0, B4, R0 | 3 | Coma |
| 28 | 1 | 0 | intubated | F=200 µg/h; P=200 mg/h | E0, M2, B2, R0 | E1, V1, M4 | Coma |
|  | 2 | 3 | none |  | 16 | 15 | FC |
|  | 3 | 5 | none |  | 16 | 15 | FC |
| 29 | 1 | 1 | intubated | F=200 µg/h; P=150 mg/h | E0, M0, B2, R0 | 3 | Coma |
| 30 | 1 | 6 | intubated | F=250 µg/h; MI=15 mg/h | E0, M2, B4, R0 | E1, V1, M4 | Coma |
| 31 | 1 | 2 | intubated | P=90 mg/h; RE=1000 µg/h | E2, M2, B4, R0 | E3, V1, M4 | UWS |
|  | 2 | 3 | none |  | 16 | E4, V4, M6 | CS |
| 32 | 1 | 0 | intubated | F=200 µg/h; P=100 mg/h | E0, M0, B4, R0 | 3 | Coma |
|  | 2 | 1 | none |  | 16 | 15 | FC |
| 33 | 1 | 0 | intubated | F=250 µg/h; P=220 mg/h | E0, M0, B4, R0 | 3 | Coma |
|  | 2 | 3 | intubated | RE=450 µg/h | E0, M0, B4, R1 | 3 | Coma |
|  | 3 | 7 | intubated |  | E0, M2, B4, R1 | E1, V1, M4 | UWS |
| 34 | 1 | 16 | intubated |  | E1, M0, B4, R1 | E2, V1, M1 | UWS |
| 35 | 1 | 4 | intubated | P=100 mg/h; RE=500 µg/h | E0, M2, B4, R0 | E1, V1, M4 | Coma |
|  | 2 | 8 | tracheostomy |  | E3, M4, B4, R1 | E3, V1, M6 | MCS+ |
|  | 3 | 12 | tracheostomy |  | E4, M4, B4, R1 | E4, V1, M6 | eMCS |
| 36 | 1 | 5 | intubated | RE=500 µg/h | E3, M2, B4, R1 | E4, V1, M4 | UWS |
|  | 2 | 10 | none |  | E3, M4, B4, R4 | E4, V1, M6 | MCS+ |
|  | 3 | 27 | intubated |  | E0, M0, B2, R1 | 3 | Coma |
| 37 | 1 | 12 | intubated | F=50 µg/h; P=160 mg/h | E0, M0, B4, R1 | 3 | Coma |
| 38 | 1 | 6 | intubated | F=700 µg/h; MI=50 mg/h | E0, M0, B4, R0 | 3 | Coma |
|  | 2 | 17 | tracheostomy |  | E1, M2, B4, R1 | E2, V1, M4 | UWS |
|  | 3 | 19 | tracheostomy |  | E0, M2, B4, R1 | E1, V1, M4 | Coma |
| 39 | 1 | 1 | intubated | F=400 µg/h; P=350 mg/h | E0, M0, B2, R0 | 3 | Coma |
| 40 | 1 | 1 | intubated | F=200 µg/h; P=200 mg/h | E0, M0, B4, R0 | 3 | Coma |
|  | 2 | 2 | intubated | F=25 µg/h | E4, M2, B4, R0 | E4, V1, M4 | MCS- |
| 41 | 1 | 1 | intubated | F=300 µg/h; P=160 mg/h | E0, M0, B4, R0 | 3 | Coma |
|  | 2 | 2 | intubated | RE=1250 µg/h | E2, M4, B4, R0 | E3, V1, M6 | MCS+ |
| 42 | 1 | 1 | intubated | F=300 µg/h; P=160 mg/h | E0, M0, B4, R0 | 3 | Coma |
|  | 2 | 10 | intubated |  | E2, M2, B4, R1 | E3, V1, M4 | UWS |
|  | 3 | 13 | tracheostomy |  | E4, M4, B4, R1 | E4, V1, M6 | eMCS |
| 43 | 1 | 1 | intubated | F=200 µg/h; P=280 mg/h | E0, M0, B2, R0 | 3 | Coma |
|  | 2 | 8 | intubated |  | E0, M2, B4, R1 | E1, V1, M4 | Coma |
| 44 | 1 | 0 | intubated | F=200 µg/h; P=70 mg/h | E0, M0, B4, R0 | 3 | Coma |
| 45 | 1 | 1 | intubated | F=100 µg/h; P=70 mg/h | E0, M0, B2, R0 | 3 | Coma |
| 46 | 1 | 2 | intubated | P=300 mg/h; RE=750 µg/h | E0, M0, B4, R0 | 3 | Coma |
|  | 2 | 11 | tracheostomy |  | E0, M2, B2, R1 | E1, V1, M4 | Coma |
|  | 3 | 13 | tracheostomy |  | E0, M2, B4, R1 | E1, V1, M4 | Coma |
| 47 | 1 | 0 | intubated | F=300 µg/h; P=200 mg/h | E0, M0, B4, R0 | 3 | Coma |
|  | 2 | 3 | intubated | RE=500 µg/h | E0, M2, B4, R0 | E1, V1, M3 | Coma |
| 48 | 1 | 22 | intubated | F=750 µg/h; MI=65 mg/h | E0, M0, B4, R0 | 3 | Coma |
|  | 2 | 27 | tracheostomy |  | E1, M0, B2, R1 | E2, V1, M1 | UWS |
| 49 | 1 | 1 | intubated | P=150 mg/h; RE=1000 µg/h | E0, M0, B0, R0 | 3 | Coma |
|  | 2 | 10 | intubated |  | E0, M2, B0, R1 | E1, V1, M4 | Coma |
|  | 3 | 13 | tracheostomy |  | E0, M2, B2, R1 | E1, V1, M4 | MCS- |
|  | 4 | 24 | tracheostomy |  | E2, M2, B4, R1 | E3, V1, M4 | MCS- |
| 50 | 1 | 1 | intubated | F=400 µg/h; P=200 mg/h | E0, M0, B2, R0 | 3 | Coma |
|  | 2 | 3 | none |  | 16 | 15 | FC |
| 51 | 1 | 5 | intubated | P=80 mg/h; RE=500 µg/h | E0, M2, B2, R1 | E1, V1, M4 | Coma |
| 52 | 1 | 1 | intubated | F=200 µg/h; P=120 mg/h | E0, M0, B2, R0 | 3 | Coma |
|  | 2 | 2 | intubated | RE=100 µg/h | E2, M2, B4, R1 | E3, V1, M4 | UWS |
| 53 | 1 | 6 | intubated | P=150 µg/h; RE=750 µg/h | E0, M2, B2, R1 | E1, V1, M4 | Coma |
| 54 | 1 | 1 | intubated | F=400 µg/h; MI=40 mg/h; P=350 mg/h | E0, M0, B2, R0 | 3 | Coma |
| 55 | 1 | 1 | intubated | F=200 µg/h; P=190 mg/h | E0, M0, B2, R0 | 3 | Coma |
|  | 2 | 3 | none |  | E2, M4, B4, R4 | E3, V2, M6 | MCS+ |
|  | 3 | 5 | none |  | E2, M4, B4, R4 | E3, V4, M6 | eMCS |
| 56 | 1 | 1 | intubated | F=200 µg/h; P=100 mg/h | E0, M0, B4, R0 | 3 | Coma |
|  | 2 | 2 | intubated |  | E3, M0, B4, R1 | E3, V1, M1 | MCS- |
|  | 3 | 3 | none |  | 16 | E4, V1, M6 | eMCS |
| 57 | 1 | 6 | intubated | F=750 µg/h; MI=87 mg/h; P=116 mg/h | E0, M0, B2, R0 | 3 | Coma |
| 58 | 1 | 2 | intubated | F=350 µg/h; MI=45 mg/h; T=300 mg/h | E0, M0, B0, R0 | 3 | Coma |
|  | 2 | 22 | tracheostomy |  | E2, M2, B2, R1 | E3, V1, M4 | UWS |
|  | 3 | 24 | tracheostomy |  | E2, M3, B4, R1 | E3, V1, M5 | MCS- |
| 59 | 1 | 1 | intubated | F=250 µg/h; P=250 mg/h | E0, M0, B2, R0 | 3 | Coma |
| 60 | 1 | 1 | intubated | P=200 mg/h; RE=600 µg/h | E0, M0, B4, R0 | 3 | Coma |
|  | 2 | 3 | none |  | 16 | E4, V4, M6 | CS |
|  | 3 | 9 | none |  | 16 | 15 | FC |
| 61 | 1 | 1 | intubated | P=150 mg/h; RE=1000 µg/h | E0, M0, B2, R0 | 3 | Coma |
|  | 2 | 10 | intubated | F=200 µg/h; MI=20 mg/h | E0, M0, B4, R1 | 3 | Coma |
|  | 3 | 13 | tracheostomy | F=150 µg/h; MI=1.8 ml/time | E0, M2, B4, R1 | E1, V1, M4 | Coma |
|  | 4 | 17 | tracheostomy |  | E0, M0, B4, R1 | 3 | Coma |
| 62 | 1 | 3 | intubated | F=400 µg/h; MI=20 mg/h | E0, M0, B4, R1 | 3 | Coma |
|  | 2 | 11 | intubated |  | E0, M0, B4, R0 | 3 | Coma |
|  | 3 | 18 | tracheostomy |  | E0, M0, B4, R1 | 3 | Coma |
| 63 | 1 | 1 | intubated | P=200 mg/h; RE=600 µg/h | E0, M0, B4, R0 | 3 | Coma |
|  | 2 | 13 | tracheostomy | RE=650 µg/h | E4, M4, B4, R1 | E4, V1, M6 | MCS+ |
| 64 | 1 | 1 | intubated | P=50 mg/h; RE=250 µg/h | E0, M2, B4, R0 | E1, V1, M4 | Coma |
| 65 | 1 | 4 | intubated | P=100 mg/h; RE=750 µg/h | E0, M2, B4, R1 | E1, V1, M4 | Coma |
|  | 2 | 5 | intubated | RE=750 µg/h | E1, M2, B4, R1 | E2, V1, M4 | UWS |
|  | 3 | 11 | intubated | RE=230 µg/h | E3, M3, B4, R1 | E3, V1, M5 | MCS- |
| 66 | 1 | 1 | intubated | P=150 mg/h; RE=1000 µg/h | E0, M0, B4, R0 | 3 | Coma |
|  | 2 | 8 | intubated | RE=200 µg/h | E0, M2, B4, R1 | E1, V1, M4 | Coma |
|  | 3 | 11 | tracheostomy |  | E0, M2, B4, R1 | E1, V1, M4 | Coma |
| 67 | 1 | 1 | intubated | F=200 µg/h; P=170 mg/h | E0, M0, B4, R0 | 3 | Coma |
|  | 2 | 2 | intubated |  | E2, M2, B4, R0 | E2, V1, M4 | UWS |
|  | 3 | 3 | intubated |  | E0, M0, B4, R0 | 3 | Coma |
| 68 | 1 | 2 | intubated | F=50 µg/h; P=60 mg/h | E0, M0, B4, R0 | 3 | Coma |
|  | 2 | 4 | none |  | E4, M4, B4, R4 | E4, V4, M6 | CS |
| 69 | 1 | 15 | tracheostomy | F=200 µg/h; MI=12.5 mg/h | E1, M0, B4 R0 | E2, V1, M1 | UWS |
|  | 2 | 17 | tracheostomy |  | E0, M2, B4, R1 | E1, V1, M4 | Coma |
| 70 | 1 | 4 | intubated | F=400 µg/h; MI=40 mg/h; T=100 mg/h | E0, M0, B2, R0 | 3 | Coma |
|  | 2 | 21 | tracheostomy |  | E0, M0, B4, R1 | 3 | Coma |
|  | 3 | 24 | tracheostomy |  | E0, M0, B4, R1 | 3 | Coma |
| 71 | 1 | 2 | intubated | P=250 mg/h; RE=1250 µg/h | E0, M0, B4, R0 | 3 | Coma |
| 72 | 1 | 4 | intubated | F=500 µg/h; MI=50 mg/h; T=250 mg/h | E0, M0, B0, R0 | 3 | Coma |
| 73 | 1 | 0 | intubated | F=200 µg/h; P=130 mg/h | E0, M0, B2, R0 | 3 | Coma |
|  | 2 | 1 | intubated | RE=750 µg/h | E3, M0, B4, R0 | E3, V1, M1 | UWS |
|  | 3 | 2 | none |  | E3, M0, B4, R4 | E3, V1, M1 | MCS- |
| 74 | 1 | 1 | intubated | P=180 mg/h; RE=750 µg/h | E0, M0, B4, R0 | 3 | Coma |
|  | 2 | 5 | intubated | RE=350 ml/time | E3, M2, B4, R1 | E4, V1, M4 | UWS |
| 75 | 1 | 9 | intubated | P=250 mg/h; RE=1250 µg/h | E0, M2, B4, R1 | E1, V1, M4 | Coma |
| 76 | 1 | 1 | intubated | F=200 µg/h; P=150 mg/h | E0, M0, B2, R0 | 3 | Coma |
| 77 | 1 | 6 | intubated | F=400 µg/h; MI=30 mg/h | E0, M0, B0, R0 | 3 | Coma |
| 78 | 1 | 4 | intubated | RE=600 µg/h | E0, M0, B2, R1 | 3 | Coma |
| 79 | 1 | 1 | intubated | F=200 µg/h; P=140 mg/h | E0, M0, B4, R0 | 3 | Coma |
|  | 2 | 3 | none |  | 16 | 15 | FC |
|  | 3 | 7 | none |  | 16 | 15 | FC |
| 80 | 1 | 1 | intubated | F=400 µg/h; P=180 mg/h | E0, M0, B2, R0 | 3 | Coma |
|  | 2 | 3 | intubated | RE=300 µg/h | E2, M4, B4, R0 | E3, V1, M6 | MCS+ |
| 81 | 1 | 2 | intubated | F=200 µg/h; P=200 mg/h | E0, M0, B2, R0 | 3 | Coma |
| 82 | 1 | 2 | intubated | P=150 mg/h; RE=750 µg/h | E0, M0, B4, R0 | 3 | Coma |
|  | 2 | 9 | tracheostomy |  | E3, M2, B4, R1 | E4, V1, M4 | UWS |
|  | 3 | 11 | tracheostomy |  | E3, M2, B4, R1 | E4, V1, M4 | UWS |
|  | 4 | 16 | tracheostomy |  | E3, M2, B4, R1 | E4, V1, M4 | MCS- |
| 83 | 1 | 0 | intubated | F=200 µg/h; P=80 mg/h | E0, M0, B2, R0 | 3 | Coma |
|  | 2 | 1 | intubated |  | E2, M0, B2, R1 | E2, V1, M1 | UWS |
| 84 | 1 | 1 | intubated | F=500 µg/h; T=150 mg/h | E0, M0, B2, R0 | 3 | Coma |
| 85 | 1 | 3 | intubated | F=500 µg/h; MI=50 mg/h | E0, M0, B4, R0 | 3 | Coma |
|  | 2 | 14 | tracheostomy |  | E2, M4, B4, R1 | E3, V1, M6 | MCS+ |
|  | 3 | 16 | tracheostomy |  | E4, M4, B4, R1 | E4, V1, M6 | eMCS |
| 86 | 1 | 0 | intubated | F=300 µg/h; P=300 mg/h | E0, M0, B2, R0 | 3 | Coma |
| 87 | 1 | 2 | intubated | P=130 mg/h; RE=650 µg/h | E0, M2, B4, R0 | E1, V1, M4 | Coma |
| 88 | 1 | 5 | intubated | RE=1500 µg/h | E2, M2, B4, R1 | E3, V1, M4 | MCS- |
|  | 2 | 8 | tracheostomy |  | E2, M2, B4, R1 | E3, V1, M4 | UWS |
| 89 | 1 | 0 | intubated | F=200 µg/h; P=180 mg/h | E0, M0, B0, R0 | 3 | Coma |
| 90 | 1 | 4 | intubated | F=300 µg/h; MI=30 mg/h | E0, M0, B2, R0 | 3 | Coma |
|  | 2 | 29 | tracheostomy | MI=5 mg injection. | E0, M0, B4, R1 | 3 | Coma |
| 91 | 1 | 1 | intubated | P=100 mg/h; RE=250 µg/h | E0, M0, B4, R1 | 3 | Coma |
|  | 2 | 6 | intubated | F=150 µg/h; MI=25 mg/h | E0, M0, B4, R0 | 3 | Coma |
| Abbreviations: B=Brainstem; CS=Confusional State; E=Eye response; eMCS=emerged from MCS; F=Fentanyl; FC=Fully Conscious State, FOUR=Four Outline of UnResponsiveness Score; GCS=Glasgow Coma Scale; LIS=Locked-in Syndrome; M=Motor response; MCS=Minimally Conscious State (minus “-“/plus “+”); MI=Midazolam; P=Propofol; R=Respiration; RE=Remifentanil; T=Thiopental; UWS=Unresponsive Wakefulness Syndrome; V=Verbal response.  *Concentrations solutions for the sedative drugs are as follows: Propofol 20 mg/ml, Fentanyl 50 µg/ml, Remifentanil 50 µg/ml, Midazolam 5mg/ml, Thiopental 25 mg/ml. | | | | | | | |

| **Supplemental Table 3: Individualized Pupillometry Results** | | | | | | | | | | | | | | | | |
| --- | --- | --- | --- | --- | --- | --- | --- | --- | --- | --- | --- | --- | --- | --- | --- | --- |
| **ID** | **Clinical**  **Examination** | **Consciousness**  **Level** | **Mirror**  **(1/1) ^a^** | **Auditory**  **(2/3) ^a^** | **MA-m**  **(3/5) ^a^** | **MA-m**  **(4/5) ^a^** | **MA-h**  **(3/5)** | **MA-h**  **(4/5)** | **Either MA ^b^**  **(3/5)** | **Both MA ^c^**  **(3/5)** | **Either MA**  **(4/5)** | **Both MA**  **(4/5)** | **Mirror**  **Record ID ^d,e^** | **Auditory**  **Record ID^e^** | **MA-m**  **Record ID^e^** | **MA-h**  **Record ID^e^** |
| 1 | 1 | Coma | no | no | no | no | no | no | no | no | no | no |  |  |  |  |
|  | 2 | Coma | no | no | no | no | no | no | no | no | no | no |  |  |  |  |
| 2 | 1 | LIS | no | no | yes | no | yes | no | yes | yes | no | no |  |  |  |  |
| 3 | 1 | Coma | yes | no | no | no | no | no | no | no | no | no |  |  |  |  |
|  | 2 | Coma | no | yes | no | no | no | no | no | no | no | no |  |  |  |  |
|  | 3 | UWS | no | no | no | no | yes | yes | yes | no | yes | no |  |  |  |  |
| 4 | 1 | Coma | no | no | no | no | no | no | no | no | no | no |  |  |  |  |
|  | 2 | MCS+ | no | no | no | no | no | no | no | no | no | no |  |  |  |  |
|  | 3 | MCS+ | yes | yes | yes | no | yes | no | yes | yes | no | no |  |  |  |  |
| 5 | 1 | Coma | no | yes | no | no | no | no | no | no | no | no |  |  |  |  |
|  | 2 | CS | no | no | no | no | no | no | no | no | no | no |  |  |  |  |
|  | 3 | FC | no | yes | yes | yes | ND | ND | yes | no | yes | no |  |  |  |  |
| 6 | 1 | MCS- | no | no | yes | yes | yes | no | yes | yes | yes | no |  |  |  |  |
|  | 2 | FC | yes | yes | yes | yes | no | no | yes | no | yes | no |  |  |  |  |
| 7 | 1 | Coma | no | no | no | no | no | no | no | no | no | no |  |  |  |  |
| 8 | 1 | Coma | no | 2 | no | no | no | no | no | no | no | no |  |  |  |  |
| 9 | 1 | Coma | no | yes | no | no | yes | no | yes | no | no | no |  |  |  |  |
| 10 | 1 | Coma | yes | yes | no | no | no | no | no | no | no | no |  |  |  |  |
| 11 | 1 | Coma | no | yes | no | no | no | no | no | no | no | no |  |  |  |  |
|  | 2 | FC | no | yes | no | no | ND | ND | no | no | no | no |  |  |  |  |
| 12 | 1 | Coma | no | yes | no | no | no | no | no | no | no | no |  |  |  |  |
|  | 2 | MCS+ | no | yes | no | no | no | no | no | no | no | no |  |  |  |  |
| 13 | 1 | Coma | no | yes | no | no | no | no | no | no | no | no |  |  |  |  |
|  | 2 | CS | no | no | no | no | ND | ND | no | no | no | no |  |  |  |  |
| 14 | 1 | Coma | no | yes | no | no | no | no | no | no | no | no |  |  |  |  |
|  | 4 | FC | no | no | no | no | no | no | no | no | no | no |  |  |  |  |
| 15 | 1 | Coma | no | yes | no | no | no | no | no | no | no | no |  |  |  |  |
|  | 2 | FC | yes | yes | no | no | yes | no | yes | no | no | no |  |  |  |  |
| 16 | 1 | MCS- | no | no | yes | no | no | no | yes | no | no | no |  |  |  |  |
| 17 | 1 | Coma | no | no | no | no | no | no | no | no | no | no |  |  |  |  |
| 18 | 1 | Coma | no | yes | yes | no | no | no | yes | no | no | no |  |  |  |  |
|  | 2 | FC | yes | no | yes | no | no | no | yes | no | no | no |  |  |  |  |
| 19 | 1 | Coma | no | yes | no | no | no | no | no | no | no | no |  |  |  |  |
| 20 | 1 | Coma | no | no | no | no | yes | no | yes | no | no | no |  |  |  |  |
|  | 2 | UWS | no | yes | yes | yes | yes | yes | yes | yes | yes | yes |  |  |  |  |
|  | 3 | UWS | no | no | yes | yes | yes | no | yes | yes | yes | no |  |  |  |  |
|  | 4 | UWS | ND | ND | no | no | no | no | no | no | no | no |  |  |  |  |
| 21 | 1 | Coma | no | yes | no | no | no | no | no | no | no | no |  |  |  |  |
| 22 | 1 | Coma | no | no | no | no | no | no | no | no | no | no |  |  |  |  |
|  | 2 | MCS+ | no | no | yes | no | ND | ND | yes | no | no | no |  |  |  |  |
| 23 | 1 | Coma | no | no | ND | ND | ND | ND | ND | ND | ND | ND |  |  |  |  |
|  | 2 | UWS | no | no | no | no | no | no | no | no | no | no |  |  |  |  |
|  | 3 | UWS | no | no | no | no | no | no | no | no | no | no |  |  |  |  |
| 24 | 1 | Coma | yes | yes | no | no | no | no | no | no | no | no |  |  |  |  |
|  | 2 | CS | no | yes | yes | yes | yes | no | yes | yes | yes | no |  |  |  |  |
| 25 | 1 | Coma | no | yes | no | no | yes | yes | yes | no | yes | no |  |  |  |  |
|  | 3 | CS | no | yes | yes | yes | ND | ND | yes | no | yes | no |  |  |  |  |
| 26 | 1 | Coma | no | yes | no | no | no | no | no | no | no | no |  |  |  |  |
| 27 | 1 | Coma | yes | yes | yes | no | yes | yes | yes | yes | yes | no |  |  |  |  |
| 28 | 1 | Coma | no | yes | no | no | no | no | no | no | no | no |  |  |  |  |
|  | 2 | FC | no | no | yes | no | no | no | yes | no | no | no |  |  |  |  |
|  | 3 | FC | no | yes | no | no | no | no | no | no | no | no |  |  |  |  |
| 29 | 1 | Coma | no | no | no | no | no | no | no | no | no | no |  |  |  |  |
| 30 | 1 | Coma | no | yes | no | no | no | no | no | no | no | no |  |  |  |  |
| 31 | 1 | UWS | no | yes | no | no | no | no | no | no | no | no |  |  |  |  |
|  | 2 | CS | yes | yes | yes | yes | ND | ND | yes | no | yes | no |  |  |  |  |
| 32 | 1 | Coma | no | yes | no | no | no | no | no | no | no | no |  |  |  |  |
|  | 2 | FC | no | yes | yes | yes | no | no | yes | no | yes | no |  |  |  |  |
| 33 | 1 | Coma | no | no | no | no | no | no | no | no | no | no |  |  |  |  |
|  | 2 | Coma | no | yes | yes | no | no | no | yes | no | no | no |  |  |  |  |
|  | 3 | UWS | yes | no | no | no | yes | yes | yes | no | yes | no |  |  |  |  |
| 34 | 1 | UWS | no | no | no | no | yes | no | yes | no | no | no |  |  |  |  |
| 35 | 1 | Coma | yes | yes | yes | yes | no | no | yes | no | yes | no |  |  |  |  |
|  | 2 | MCS+ | no | yes | no | no | no | no | no | no | no | no |  |  |  |  |
|  | 3 | eMCS | no | no | yes | yes | no | no | yes | no | yes | no |  |  |  |  |
| 36 | 1 | UWS | no | yes | no | no | no | no | no | no | no | no |  |  |  |  |
|  | 2 | MCS+ | no | no | no | no | no | no | no | no | no | no |  |  |  |  |
|  | 3 | Coma | no | no | yes | no | no | no | yes | no | no | no |  |  |  |  |
| 37 | 1 | Coma | no | no | no | no | no | no | no | no | no | no |  |  |  |  |
| 38 | 1 | Coma | no | no | yes | yes | no | no | yes | no | yes | no |  |  |  |  |
|  | 2 | UWS | no | no | no | no | no | no | no | no | no | no |  |  |  |  |
|  | 3 | Coma | no | no | no | no | no | no | no | no | no | no |  |  |  |  |
| 39 | 1 | Coma | yes | yes | yes | no | no | no | yes | no | no | no |  |  |  |  |
| 40 | 1 | Coma | yes | yes | no | no | yes | no | yes | no | no | no |  |  |  |  |
|  | 2 | MCS- | no | yes | no | no | no | no | no | no | no | no |  |  |  |  |
| 41 | 1 | Coma | no | yes | no | no | no | no | no | no | no | no |  |  |  |  |
|  | 2 | MCS+ | no | no | yes | no | no | no | yes | no | no | no |  |  |  |  |
| 42 | 1 | Coma | no | yes | no | no | no | no | no | no | no | no |  |  |  |  |
|  | 2 | UWS | no | no | no | no | no | no | no | no | no | no |  |  |  |  |
|  | 3 | eMCS | no | yes | no | no | yes | no | yes | no | no | no |  |  |  |  |
| 43 | 1 | Coma | no | no | no | no | no | no | no | no | no | no |  |  |  |  |
|  | 2 | Coma | no | no | no | no | no | no | no | no | no | no |  |  |  |  |
| 44 | 1 | Coma | no | yes | no | no | no | no | no | no | no | no |  |  |  |  |
| 45 | 1 | Coma | yes | no | no | no | no | no | no | no | no | no |  |  |  |  |
| 46 | 1 | Coma | no | no | no | no | no | no | no | no | no | no |  |  |  |  |
|  | 2 | Coma | no | no | no | no | no | no | no | no | no | no |  |  |  |  |
|  | 3 | Coma | no | no | no | no | no | no | no | no | no | no |  |  |  |  |
| 47 | 1 | Coma | yes | no | no | no | no | no | no | no | no | no |  |  |  |  |
|  | 2 | Coma | no | yes | yes | yes | no | no | yes | no | yes | no |  |  |  |  |
| 48 | 1 | Coma | no | yes | no | no | no | no | no | no | no | no |  |  |  |  |
|  | 2 | UWS | no | no | no | no | no | no | no | no | no | no |  |  |  |  |
| 49 | 1 | Coma | no | yes | no | no | no | no | no | no | no | no |  |  |  |  |
|  | 2 | Coma | no | yes | no | no | no | no | no | no | no | no |  |  |  |  |
|  | 3 | MCS- | no | yes | no | no | no | no | no | no | no | no |  |  |  |  |
|  | 4 | MCS- | no | yes | no | no | no | no | no | no | no | no |  |  |  |  |
| 50 | 1 | Coma | no | no | no | no | no | no | no | no | no | no |  |  |  |  |
|  | 2 | FC | yes | yes | no | no | yes | no | yes | no | no | no |  |  |  |  |
| 51 | 1 | Coma | no | no | no | no | no | no | no | no | no | no |  |  |  |  |
| 52 | 1 | Coma | no | yes | no | no | no | no | no | no | no | no |  |  |  |  |
|  | 2 | UWS | no | yes | yes | no | no | no | yes | no | no | no |  |  |  |  |
| 53 | 1 | Coma | no | no | no | no | yes | yes | yes | no | yes | no |  |  |  |  |
| 54 | 1 | Coma | no | yes | no | no | yes | no | yes | no | no | no |  |  |  |  |
| 55 | 1 | Coma | no | yes | no | no | no | no | no | no | no | no |  |  |  |  |
|  | 2 | MCS+ | no | yes | yes | yes | no | no | yes | no | yes | no |  |  |  |  |
|  | 3 | eMCS | no | no | yes | no | yes | yes | yes | yes | yes | no |  |  |  |  |
| 56 | 1 | Coma | no | yes | no | no | no | no | no | no | no | no |  |  |  |  |
|  | 2 | MCS- | no | no | no | no | yes | no | yes | no | no | no |  |  |  |  |
|  | 3 | eMCS | no | yes | no | no | ND | ND | no | no | no | no |  |  |  |  |
| 57 | 1 | Coma | yes | no | no | no | yes | yes | yes | no | yes | no |  |  |  |  |
| 58 | 1 | Coma | no | yes | no | no | no | no | no | no | no | no |  |  |  |  |
|  | 2 | UWS | no | no | no | no | yes | no | yes | no | no | no |  |  |  |  |
|  | 3 | MCS- | yes | no | no | no | yes | no | yes | no | no | no |  |  |  |  |
| 59 | 1 | Coma | no | yes | no | no | no | no | no | no | no | no |  |  |  |  |
| 60 | 1 | Coma | yes | yes | no | no | no | no | no | no | no | no |  |  |  |  |
|  | 2 | CS | no | yes | yes | yes | yes | yes | yes | yes | yes | yes |  |  |  |  |
|  | 3 | FC | no | yes | yes | yes | yes | yes | yes | yes | yes | yes |  |  |  |  |
| 61 | 1 | Coma | no | no | no | no | no | no | no | no | no | no |  |  |  |  |
|  | 2 | Coma | no | no | no | no | no | no | no | no | no | no |  |  |  |  |
|  | 3 | Coma | no | no | no | no | yes | no | yes | no | no | no |  |  |  |  |
|  | 4 | Coma | no | yes | no | no | ND | ND | no | no | no | no |  |  |  |  |
| 62 | 1 | Coma | yes | no | no | no | no | no | no | no | no | no |  |  |  |  |
|  | 2 | Coma | no | no | yes | no | no | no | yes | no | no | no |  |  |  |  |
|  | 3 | Coma | no | no | yes | no | yes | yes | yes | yes | yes | no |  |  |  |  |
| 63 | 1 | Coma | no | yes | yes | no | no | no | yes | no | no | no |  |  |  |  |
|  | 2 | MCS+ | no | yes | yes | no | no | no | yes | no | no | no |  |  |  |  |
| 64 | 1 | Coma | no | yes | yes | no | no | no | yes | no | no | no |  |  |  |  |
| 65 | 1 | Coma | no | yes | no | no | no | no | no | no | no | no |  |  |  |  |
|  | 2 | UWS | yes | no | no | no | no | no | no | no | no | no |  |  |  |  |
|  | 3 | MCS- | no | no | yes | no | yes | no | yes | yes | no | no |  |  |  |  |
| 66 | 1 | Coma | yes | yes | yes | no | no | no | yes | no | no | no |  |  |  |  |
|  | 2 | Coma | no | no | no | no | yes | no | yes | no | no | no |  |  |  |  |
|  | 3 | Coma | no | yes | no | no | no | no | no | no | no | no |  |  |  |  |
| 67 | 1 | Coma | yes | no | no | no | no | no | no | no | no | no |  |  |  |  |
|  | 2 | UWS | no | yes | no | no | no | no | no | no | no | no |  |  |  |  |
|  | 3 | Coma | yes | no | yes | no | no | no | yes | no | no | no |  |  |  |  |
| 68 | 1 | Coma | no | yes | no | no | yes | no | yes | no | no | no |  |  |  |  |
|  | 2 | CS | yes | yes | yes | yes | yes | no | yes | yes | yes | no |  |  |  |  |
| 69 | 1 | UWS | no | no | no | no | no | no | no | no | no | no |  |  |  |  |
|  | 2 | Coma | yes | yes | no | no | no | no | no | no | no | no |  |  |  |  |
| 70 | 1 | Coma | no | no | no | no | no | no | no | no | no | no |  |  |  |  |
|  | 2 | Coma | no | no | yes | no | no | no | yes | no | no | no |  |  |  |  |
|  | 3 | Coma | no | no | yes | yes | yes | no | yes | yes | yes | no |  |  |  |  |
| 71 | 1 | Coma | no | yes | no | no | no | no | no | no | no | no |  |  |  |  |
| 72 | 1 | Coma | yes | no | no | no | no | no | no | no | no | no |  |  |  |  |
| 73 | 1 | Coma | no | yes | no | no | no | no | no | no | no | no |  |  |  |  |
|  | 2 | UWS | no | yes | no | no | yes | no | yes | no | no | no |  |  |  |  |
|  | 3 | MCS- | no | no | no | no | yes | yes | yes | no | yes | no |  |  |  |  |
| 74 | 1 | Coma | no | yes | no | no | no | no | no | no | no | no |  |  |  |  |
|  | 2 | UWS | no | no | no | no | yes | yes | yes | no | yes | no |  |  |  |  |
| 75 | 1 | Coma | no | yes | no | no | no | no | no | no | no | no |  |  |  |  |
| 76 | 1 | Coma | no | yes | no | no | no | no | no | no | no | no |  |  |  |  |
| 77 | 1 | Coma | no | no | no | no | no | no | no | no | no | no |  |  |  |  |
| 78 | 1 | Coma | no | no | no | no | no | no | no | no | no | no |  |  |  |  |
| 79 | 1 | Coma | no | yes | no | no | no | no | no | no | no | no |  |  |  |  |
|  | 2 | FC | no | no | no | no | yes | no | yes | no | no | no |  |  |  |  |
|  | 3 | FC | no | no | yes | yes | yes | no | yes | yes | yes | no |  |  |  |  |
| 80 | 1 | Coma | no | no | no | no | ND | ND | no | no | no | no |  |  |  |  |
|  | 2 | MCS+ | no | no | yes | yes | yes | yes | yes | yes | yes | yes |  |  |  |  |
| 81 | 1 | Coma | yes | yes | no | no | no | no | no | no | no | no |  |  |  |  |
| 82 | 1 | Coma | no | yes | yes | yes | yes | no | yes | yes | yes | no |  |  |  |  |
|  | 2 | UWS | no | no | no | no | no | no | no | no | no | no |  |  |  |  |
|  | 3 | UWS | no | no | no | no | no | no | no | no | no | no |  |  |  |  |
|  | 4 | MCS- | no | yes | no | no | no | no | no | no | no | no |  |  |  |  |
| 83 | 1 | Coma | yes | no | no | no | no | no | no | no | no | no |  |  |  |  |
|  | 2 | UWS | no | yes | yes | yes | yes | no | yes | yes | yes | no |  |  |  |  |
| 84 | 1 | Coma | no | no | no | no | no | no | no | no | no | no |  |  |  |  |
| 85 | 1 | Coma | yes | no | no | no | no | no | no | no | no | no |  |  |  |  |
|  | 2 | MCS+ | no | no | no | no | no | no | no | no | no | no |  |  |  |  |
|  | 3 | eMCS | yes | no | no | no | no | no | no | no | no | no |  |  |  |  |
| 86 | 1 | Coma | no | no | no | no | no | no | no | no | no | no |  |  |  |  |
| 87 | 1 | Coma | no | yes | yes | no | yes | no | yes | yes | no | no |  |  |  |  |
| 88 | 1 | MCS- | no | no | no | no | no | no | no | no | no | no |  |  |  |  |
|  | 2 | UWS | no | yes | no | no | no | no | no | no | no | no |  |  |  |  |
| 89 | 1 | Coma | no | no | no | no | no | no | no | no | no | no |  |  |  |  |
| 90 | 1 | Coma | yes | yes | no | no | no | no | no | no | no | no |  |  |  |  |
|  | 2 | Coma | no | yes | yes | no | no | no | yes | no | no | no |  |  |  |  |
| 91 | 1 | Coma | no | yes | no | no | no | no | no | no | no | no |  |  |  |  |
|  | 2 | Coma | no | yes | no | no | yes | no | yes | no | no | no |  |  |  |  |
| Abbreviations: CS=Confusional State; eMCS=emerged from MCS; FC=Fully Conscious State; LIS=Locked-in Syndrome; MA-m=Moderate-level Mental Arithmetic; MA-h=Hard-level Mental Arithmetic; MCS=Minimally Conscious State (minus “-“/plus “+”); UWS=Unresponsive Wakefulness Syndrome.  ^a^ The criteria for success vary depending on the paradigm and include: (1) one pupillary dilation in response to mirror reflection, (2) at least two pupillary dilations in response to three different auditory stimuli, or (3) at least three or four pupillary dilations in response to five mental arithmetic tasks. A successful outcome is indicated by "yes."  ^b^ Success was achieved if the participant demonstrated at least three or four pupillary dilations during *either* the moderate- OR hard-level mental arithmetic tasks.  ^c^ Success was achieved if the participant demonstrated at least three or four pupillary dilations during *both* the moderate- AND hard-level mental arithmetic tasks.  ^d^ The "Record ID" refers to the specific instance of data recording, which is visualized in a separate document titled “Record ID – Pupillary Graphs.”  ^e^ IDs were removed to comply with MedRxiv instructions. | | | | | | | | | | | | | | | | |

| Supplemental Table 4. Average Pupillary Dilation (mm). | | | | |
| --- | --- | --- | --- | --- |
|  | Mirror | Auditory | Moderate* | Hard* |
| Coma | 0.21 | 1.70 | 1.54 | 1.45 |
| UWS | 0.13 | 1.13 | 1.46 | 2.04 |
| MCS | 0.00 | 1.45 | 2.14 | 1.95 |
| eMCS | 0.26 | 1.73 | 3.00 | 2.55 |
| Controls | 0.53 | 1.5 | 3.81 | 3.5 |
| Abbreviations: UWS=Unresponsive Wakefulness syndrome, MCS=Minimally Conscious State, eMCS=Emerged from MCS. | | | | |

| Supplemental Table 5. Stratified group comparisons - successful pupillary dilations in response to passive and active paradigms. | | | | |
| --- | --- | --- | --- | --- |
| OHCA Patients (n=45) | **≤UWS, n (%)** | **≥MCS, n (%)** | **Odds ratio (CI 95%)** | **P-value** |
| Mirror^a^ | 11 (20.8) | 5 (15.6) | 0.71 (0.17- 2.53) | 0.775 |
| Auditory^b^ | 36 (67.9) | 15 (50.0) | 0.48 (0.17 - 1.31) | 0.159 |
| Moderate-level mental arithmetic (3/5)^c^ | 8 (15.1) | 16 (55.2) | 6.73 (2.16 - 22.77) | <0.001* |
| Hard-level mental arithmetic  (3/5) | 8 (15.3) | 13 (56.5) | 6.92 (2.05 - 25.40) | <0.001* |
| Moderate-level mental arithmetic (4/5)^d^ | 2 (3.8) | 12 (41.2) | 17.27 (3.36 - 173.93) | <0.001* |
| Hard-level mental arithmetic  (4/5) | 6 (7.8) | 2 (11.1) | 0.73 (0.07 - 4.56) | 1.000 |
| Non-Anoxic Brain Injuries (n=46) |  |  |  |  |
| Mirror | 14 (18.2) | 2 (11.1) | 0.57 (0.06 - 2.88) | 0.728 |
| Auditory | 31 (40.3) | 9 (50.0) | 1.48 (0.46 - 4.72) | 0.597 |
| Moderate-level mental arithmetic (3/5) | 16 (20.8) | 7 (38.9) | 2.40 (0.68 - 8.15) | 0.130 |
| Hard-level mental arithmetic  (3/5) | 18 (23.7) | 5 (29.4) | 1.34 (0.32 - 4.81) | 0.756 |
| Moderate-level mental arithmetic (4/5) | 6 (7.0) | 2 (11.1) | 1.47 (0.13 - 9.28) | 0.644 |
| Hard-level mental arithmetic  (4/5) | 6 (7.9) | 0 (0.0) | 0.00 (0.00 - 3.86) | 0.588 |
| Abbreviations: UWS=Unresponsive Wakefulness syndrome, MCS=Minimally Conscious State, OHCA=Out-of-Hospital Cardiac Arrest.  ^a^ Success was achieved if the participant demonstrated at least 1 of 1 pupillary dilation during a visual stimulus.  ^b^ Success was achieved if the participant demonstrated at least 2 of 3 pupillary dilations during auditory stimuli.  ^c^ Success was achieved if the participant demonstrated at least 3 of 5 pupillary dilations during mental arithmetic tasks.  ^d^ Success was achieved if the participant demonstrated at least 4 of 5 pupillary dilations during mental arithmetic tasks.  *Statistically significant | | | | |
